# Effects of Melanopic Equivalent Daylight Illuminance on Sleep Regulation and Chronotype-Specific Responses in Young Adults

**DOI:** 10.1101/2025.10.21.25338466

**Authors:** Eunji Hwang, Hyeonjin Kim, Hahyun Lee, Hyunwoo Nam, Jun-Young Lee

**Author notes:** Corresponding author. Hyunwoo Nam, Department of Neurology, Seoul Metropolitan Government-Seoul National University Boramae Medical Center, 20 Boramae-Ro 5-Gil, Dongjak-gu, Seoul, 07061, Republic of Korea. Jun-Young Lee, Department of Psychiatry, Seoul Metropolitan Government-Seoul National University Boramae Medical Center, 20 Boramae-Ro 5-Gil, Dongjak-gu, Seoul, 07061, Republic of Korea.

## Abstract

**Objectives:** Light is a key regulator of the human circadian system, yet conventional photopic illuminance does not reflect the spectral sensitivity of intrinsically photosensitive retinal ganglion cells (ipRGCs). Melanopic equivalent daylight illuminance (EDI) provides a more biologically relevant metric. We examined whether melanopic EDI better predicts real-world sleep outcomes than photopic illuminance and whether these associations differ by chronotype.

**Methods:** Fifty-nine young adults wore actigraphs for seven days to monitor light exposure and sleep. Light exposure was quantified as photopic illuminance and melanopic EDI. Chronotype was classified using the Morningness-Eveningness Questionnaire (MEQ) as non-evening type (nET) and evening type (ET). Hierarchical regression assessed the added predictive value of melanopic EDI, and linear mixed-effects models examined temporal associations and chronotype effects.

**Results:** Melanopic EDI improved the prediction of sleep outcomes across all time windows. The largest delta R2 occurred for sleep quality in the afternoon (1.55%) and at night (1.00%), deep sleep at night (0.93%), and fragmentation in the afternoon (0.51%). Nighttime exposure (01:00-03:00) was associated with poorer sleep quality in both chronotypes (nET: p=0.018 (01:00), p=0.015 (03:00); ET: p=0.043 (01:00)). Morning exposure (10:00) improved sleep quality (p=0.002) and reduced sleep fragmentation (p=0.031) in nET, whereas evening exposure (18:00–24:00) was associated with lower sleep quality (p=0.002) and greater sleep fragmentation (p=0.027) in ET.

**Conclusions:** Melanopic EDI is more sensitive to sleep than photopic illuminance. Morning light benefited nET, whereas evening light disrupted sleep in ET, supporting melanopic metrics and chronotype-based light strategies to improve sleep health.

**Statement of Significance:** Light, especially short-wavelength blue light, exerts non-visual effects on human physiology through ipRGCs that synchronize the circadian system. However, conventional light metrics, such as photopic illuminance, do not capture these spectral characteristics, limiting their ability to predict physiological outcomes. Using real-world data, this study demonstrates that melanopic EDI is a more sensitive predictor of sleep quality and structure than photopic illuminance. Temporal and chronotype-specific analyses showed that morning melanopic light improves sleep in nET, whereas evening exposure disrupts sleep in ET. These findings address a gap by demonstrating the ecological validity and chronotype-dependent impact of melanopic-sensitive light metrics. These results underscore the need to incorporate melanopic metrics and chronotype considerations into personalized sleep hygiene strategies, clinical recommendations, and public health guidelines, and point toward developing targeted interventions that leverage spectral light properties to optimize circadian and sleep health.

## Introduction

Light is the central modulator of the human circadian system, synchronizing daily rhythms of sleep–wake cycles, hormonal secretion, and metabolic processes through retinal photoreception and neural signaling to the suprachiasmatic nucleus [1–3]. Short-wavelength blue-enriched light exerts a particularly strong influence on circadian rhythm by activating intrinsically photosensitive retinal ganglion cells (ipRGCs). Through melanopsin-based phototransduction, ipRGCs regulate non-visual pathways that control circadian phase, suppress nocturnal melatonin secretion, and promote alertness [4–6]. The discovery of the central role of ipRGCs has shifted the understanding of light from being solely a visual input to being a biological regulator of physiology, highlighting the need for light metrics that capture these non-visual effects.

The biological importance of light exposure extends beyond circadian entrainment to broader domains of health. Natural daylight stabilizes internal rhythms, supports neuroendocrine balance, and contributes to mood regulation and cognitive performance [7]. In contrast, mistimed or insufficient light exposure has been implicated in a wide spectrum of health concerns. Morning light promotes circadian alignment by advancing phase and consolidating sleep, whereas evening or nighttime light delays circadian phase, suppresses melatonin, and disrupts sleep continuity [8]. Prolonged circadian misalignment, often described as ―social jetlag,‖ has been linked to decreased academic and occupational performance, heightened risk of affective and anxiety disorders, metabolic dysfunction, cardiovascular disease, and even neurodegenerative processes [9]. These findings underscore the clinical relevance of understanding how both the timing and quality of light exposure influence human physiology.

These challenges are exacerbated by modern lifestyles. In urban environments, people are exposed to limited daylight because of indoor routines, while they are surrounded by artificial lighting during the evening and night [10–12]. The widespread use of light-emitting devices such as smartphones, tablets, and LED lighting has become a significant source of short-wavelength exposure during times when the circadian system is most vulnerable. This imbalance, insufficient morning light and excessive evening illumination, contributes to circadian misalignment and associated health risks. The increasing prevalence of delayed sleep–wake phase disorder and other circadian rhythm sleep disorders is thought to reflect, in part, these environmental shifts. This chronic exposure disrupts circadian rhythms and alters neural processes in mood regulation, increasing vulnerability to metabolic, oncologic, and psychiatric disorders [13–18].

Light and sleep are influenced not only by environmental conditions but also by substantial inter-individual variability. Emerging evidence shows that sensitivity to light exposure, particularly at night, differs widely across individuals due to factors such as age, sex, chronotype, and genetic background [19]. These differences arise from both retinal mechanisms and downstream circadian pathways, leading to variability in outcomes such as melatonin suppression, circadian phase shifts, and sleep disruption. Individuals with a later circadian phase are more vulnerable to attentional lapses and neurobehavioral impairment following sleep restriction, indicating that circadian timing strongly moderates light-related effects [20, 21]. Similarly, sleep EEG characteristics show high intra-individual stability but substantial inter-individual variability, suggesting that light–sleep associations are also likely shaped by inherent individual traits [22]. These findings underscore the importance of considering individual characteristics, especially chronotype, when assessing the impact of light exposure on circadian physiology and sleep in daily life.

Despite this recognition, methodological limitations remain. Most previous studies have relied on photopic illuminance, a metric based on cone sensitivity that adequately represents visual brightness but does not capture spectral composition [23–25]. Because photopic illuminance disregards the wavelength-specific biological potency of light, it may underestimate the true circadian impact of short-wavelength exposure. To address this limitation, the Commission Internationale de l’Éclairage (CIE) recently established melanopic equivalent daylight illuminance (EDI) as a standardized measure that weights light exposure according to melanopsin sensitivity [26].

Another enduring challenge pertains to ecological validity. While this biologically relevant metric has proven useful in controlled laboratory studies and theoretical models, its application in longitudinal, real-world contexts remains limited. Thus, whether melanopic EDI provides superior predictive value for sleep outcomes compared to conventional photopic measures in the real-world is not yet established. Laboratory-based paradigms cannot fully capture the complexity, variability, and contextual determinants of real-world light exposure [27]. In real-world settings, individuals encounter dynamically fluctuating intensities and spectra, influenced by behavioral routines, occupational demands, and social activities. Students, shift workers, and office employees exhibit widely divergent patterns of light exposure that cannot be easily reproduced in laboratory environments. Recent advances in wearable, near-eye light sensors now allow continuous, ecologically valid measurement of light exposure, enabling the integration of spectral sensitivity into real-world assessments [28]. These technologies enhance translational relevance and support individualized recommendations for light and circadian health.

Chronotype, which captures intrinsic circadian phase and sleep–wake preferences, is a major source of variability in light sensitivity. By classifying individuals such as morning larks or night owls, researchers can better understand the variability in light sensitivity that arises from these differences in chronotype [29]. Melatonin regulation plays an essential role in maintaining a healthy circadian rhythm, as its production is influenced by light exposure and chronotype [30]. Understanding how chronotype, circadian rhythms, and light sensitivity interact is critical for strategies to optimize light exposure and mitigate its physiological effects [31].

This study was designed with three objectives. We evaluated whether melanopic EDI provides superior predictive value for sleep outcomes compared with conventional photopic illuminance in ecologically valid conditions. We also examined the temporal specificity of light–sleep associations by identifying windows in which exposure exerts either beneficial or disruptive effects. In addition, we further examined whether these associations are moderated by chronotype. This study aimed to advance understanding of light–sleep interactions in real-world contexts and to provide insights for personalized approaches to circadian and sleep health.

## Materials and methods

### Participants

A sample size of 60 was set to provide adequate statistical power for detecting medium-sized effects in models, consistent with prior studies on light and sleep [32]. Sixty individuals aged 19–45 years were enrolled; of these, 59 contributed at least three valid days of data and were included in the final analysis.

Exclusion criteria were severe ophthalmic conditions (e.g., blindness, glaucoma, retinal disorders), ophthalmic surgery within the past six months, diagnosed sleep disorders, use of medications affecting sleep, recent transmeridian travel, shift work, and moderate to severe symptoms of depression, anxiety, or insomnia. These criteria were defined using validated thresholds, including the Beck Depression Inventory (BDI ≥ 20), the Beck Anxiety Inventory (BAI ≥ 16) [33, 34], and the Insomnia Severity Index (ISI ≥ 22) [35].

Recruitment was conducted through advertisements distributed via online platforms, universities, and healthcare organizations. Written informed consent was obtained from all participants prior to enrollment. The study protocol was approved by the Boramae Hospital Research Ethics Review Board (IRB No. 10-2024-42).

Chronotype was determined using the Morningness–Eveningness Questionnaire (MEQ). Participants were stratified into non-evening type (nET; MEQ > 41) and evening type (ET; MEQ ≤ 41) [36]. In accordance with established criteria, the nET group comprised both intermediate and morning chronotypes.

### Data Collection

Data were collected between May 15 and August 15, 2024, to minimize potential confounding from seasonal variation. Each participant underwent continuous monitoring for seven consecutive days with individually scheduled start dates. Participants who did not complete the full 7-day protocol were retained in the analysis if at least three valid days of light and sleep data were available.

### Light and Sleep Measurements

Light exposure was measured with wearable sensors attached to the upper chest, oriented toward the eyes. Participants were instructed to wear the device continuously during waking hours and to position it near the head during sleep. The device captured melanopic EDI, photopic illuminance, RGB spectral components, and correlated color temperature at 15-second intervals. Hourly averages were derived for analysis. This type of sensor has been widely used in circadian rhythm research, providing ecologically valid data on light exposure, sleep, alertness, and circadian timing [37].

Specifically, light was recorded using a wearable melanopic EDI sensor (LYS Button, LYS Technologies Ltd., Copenhagen, Denmark). The device integrates tri-stimulus RGB and near-infrared photodiodes (spectral range: 380–1100 nm) that convert spectral irradiance into photopic and melanopic illuminance metrics. It operates across a dynamic range of 0–100,000 lx with a 15-second sampling rate and transmits data via Bluetooth to the LYS Collect mobile application. Validation studies have shown strong agreement with laboratory-grade spectroradiometers (r = 0.96–1.00; ICC = 0.90–0.98) and stable measurement bias even under high-luminance conditions (>2,500 lx), confirming the device’s reliability in real-world environments [38]. Sensors were positioned near the shirt collar to approximate corneal illumination while minimizing motion artefacts.

Sleep was assessed using the Fitbit Versa 4, wrist-worn tracker that has been widely utilized in sleep and circadian research. Device-derived sleep parameters included overall sleep score, stage-specific estimates of deep sleep duration, and restlessness, which was used as a proxy for sleep fragmentation in subsequent analyses. Sleep quality and deep sleep were analyzed as continuous measures, while sleep fragmentation was operationalized using restlessness scores derived from nocturnal movement. Daily sleep onset and offset times were automatically detected by the device and temporally aligned with light exposure data.

The Fitbit Versa 4 combines tri-axial accelerometry with optical sensors for heart rate, blood oxygen saturation (SpO₂), and skin temperature, classifying sleep stages using a proprietary algorithm. Recent validation and meta-analytic evidence indicate that Fitbit devices slightly underestimate total sleep time (by approximately 15– 20 minutes) and overestimate wake after sleep onset compared with polysomnography. However, such bias is systematic and consistent across nights and individuals, supporting their reliability for within-subject comparisons [38]. The Versa series shares the same sensing architecture and algorithmic framework as the Fitbit Inspire 3, which has been validated in Korean cohorts [40].

### Behavioral and Chronotype Assessments

Demographic and behavioral characteristics, including age, sex, employment status, educational attainment, and self-rated health, were collected as covariates. Psychological status was evaluated using validated questionnaires, including the BDI for depressive symptoms and the BAI for anxiety symptoms. Physical activity was assessed with the International Physical Activity Questionnaire, which quantifies activity frequency, intensity, and duration [41]. In addition to device-based measures, sleep quality was further evaluated using the Pittsburgh Sleep Quality Index (PSQI) [42]. Chronotype was assessed using both the Morningness–Eveningness Questionnaire (MEQ) and the Munich Chronotype Questionnaire (MCTQ) [43]. The MEQ reflects subjective morning–evening preference, while the MCTQ captures actual sleep–wake behavior based on differences between workdays and free days. Using both instruments enabled us to account for both preference and real-world behavior, providing a more comprehensive classification of chronotype for light–sleep analyses.

### Data Analysis

To assess the incremental predictive value of melanopic EDI over photopic illuminance, hierarchical regression models were applied for sleep quality, deep sleep duration, and sleep fragmentation [44, 45]. Predictors were entered sequentially to assess incremental explanatory power. Model performance was evaluated using adjusted R² and the Akaike Information Criterion (AIC) [46], with likelihood ratio tests used to assess fixed effects. Sensitivity analyses were performed after excluding outliers and high-leverage cases. Model assumptions were examined through variance inflation factors (VIFs), residual plots, and Q–Q plots [47].

Linear mixed-effects models were employed to examine temporal associations between light exposure and sleep outcomes [48, 49]. Separate models were constructed using 6-hour and 1-hour intervals. Both melanopic EDI and photopic illuminance were log-transformed to reduce skewness and heteroscedasticity [50]. Restricted maximum likelihood estimation was used for model fitting [51]. Normality and homoscedasticity assumptions were verified with the Shapiro–Wilk and Levene’s tests [52].

Group differences between ET and nET were assessed using independent two-sample t-tests for continuous variables and chi-square or Fisher’s exact tests for categorical variables [53, 54]. Assumptions were checked with the Shapiro–Wilk and Levene’s tests. Effect estimates from linear mixed-effects models are reported with SE, and error bars in figures represent ±1 SE. Statistical significance was determined from model-derived p-values (p < 0.05), not by error bar overlap with zero. All analyses were conducted in R version 4.4.2.

Analyses were conducted using fixed clock-time intervals rather than aligning to individual Tmin. This approach was chosen to examine the real-world temporal patterns of light exposure and sleep across the 24-h day, rather than circadian phase–referenced effects. As all participants underwent identical one-week monitoring, this allowed for direct comparison between chronotypes in real-life contexts.

Figures display model-derived effect estimates (β) with ±1 standard error (SE). In bar charts of ΔR², the values indicate the mean change in explained variance after adding melanopic EDI to photopic models. Statistical significance is indicated by colored markers or hatched bars (p < .05), while non-significant estimates are shown in gray. The x-axis represents either 6-hour intervals (00–06, 06–12, 12–18, 18–24) or hourly clock time (00:00– 24:00), and the y-axis indicates either Effect estimate (β) or Mean ΔR² (%), as labeled.

## Results

### Demographic and Sleep Characteristics

Participant demographic and sleep characteristics are summarized (Table 1). Based on MEQ scores, 21 individuals (35.6%) were classified as ET and 38 (64.4%) as nET. ET participants were significantly younger (23.7 ± 2.7 years) than nET participants (27.5 ± 4.9 years; p < 0.001) and had fewer years of education (p = 0.006). A larger proportion of ET participants were students.

**Table 1.** Demographic and Sleep Characteristics of Participants by Chronotype. MEQ: Morningness–Eveningness Questionnaire (scores ≥42 classified as non-Evening type; <42 classified as Evening type). PSQI: Pittsburgh Sleep Quality Index (scores ≥5 classified as poor sleep quality). BDI: Beck Depression Inventory-II (scores ≥20 indicating moderate to severe depression). Employment status values indicate the proportion of participants who were employed, unemployed, or students. Sleep metrics include overall sleep score, deep sleep duration, fragmentation, sleep onset time, and wake time (all expressed as mean and SD).

Group differences were also evident in sleep parameters. ET participants reported lower overall sleep quality (p < 0.001), shorter deep sleep duration (p < 0.001), and later sleep onset times (p = 0.006). They additionally showed higher levels of sleep fragmentation (p < 0.001). Using the PSQI cutoff of 5 to indicate poor sleep quality, the mean score for ET participants (5.4) placed them in the poor sleeper range, whereas nET participants (mean = 4.6) fell within the good sleeper range, confirming a significant group difference in subjective sleep quality.

### Superiority of Melanopic EDI in Modeling Sleep Parameters

To evaluate the incremental predictive value of melanopic EDI compared with conventional photopic illuminance, we examined the change in explained variance (ΔR²) for each sleep outcome when melanopic EDI was added to the model. Across all participants (Figure 1A–1C), the largest gain in R² for sleep quality occurred during the afternoon (ΔR² = 1.55%) and nighttime (ΔR² = 1.00%). For deep sleep duration, the greatest improvement was observed at night (ΔR² = 0.93%), whereas prediction of sleep fragmentation improved most with afternoon exposure (ΔR²= 0.51%).

**Figure. 1.**
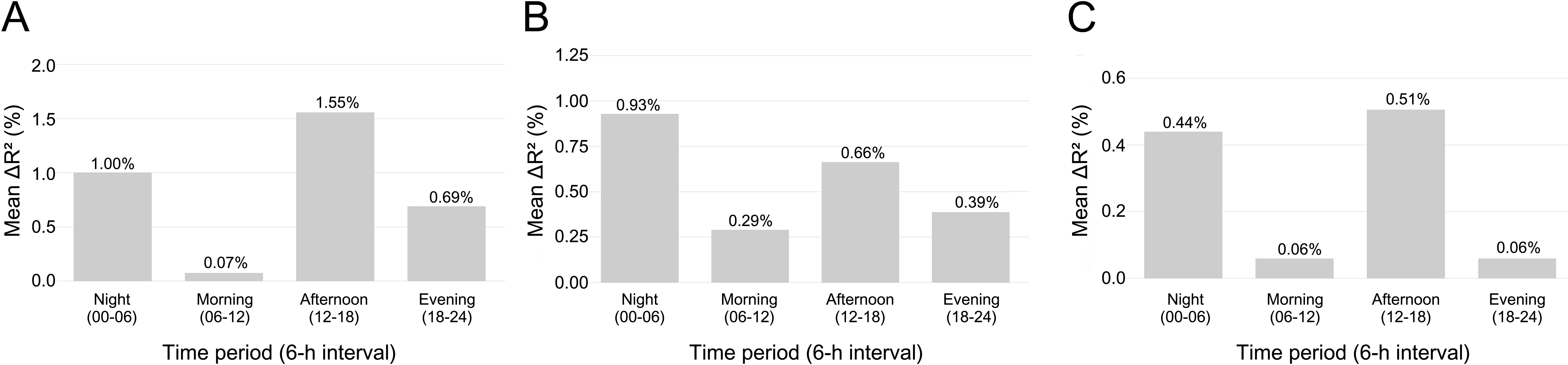
Improvement in explained variance (ΔR²) by Melanopic Equivalent Daylight Illuminance compared with photopic illuminance (6-hour interval) (A) sleep quality Score, (B) deep sleep duration, and (C) sleep fragmentation. Bars indicate mean ΔR² (%), representing the gain in explained variance after adding melanopic EDI to photopic models. Labels on each bar show the exact ΔR². Error bars are omitted by design because ΔR² is a model-comparison metric.

### Hourly Effects of Light Exposure on Sleep Parameters

While photopic illuminance also showed significant associations at limited time windows, particularly for sleep fragmentation around midday, these effects were fewer and less robust than those observed for melanopic EDI (Figure 2). Sleep quality significantly declined by melatonin-suppressing light exposures at 00:00 (β = –0.582, p = 0.022), 01:00 (β = –0.839, p = 0.001), 02:00 (β = –0.580, p = 0.035), and 03:00 (β = –0.556, p = 0.046), with additional declines at 16:00 (β = –0.866, p = 0.002) and 17:00 (β = –0.848, p = 0.006) (Figure 2A). Deep sleep duration was significantly reduced by melatonin-suppressing light exposures at 01:00 (β = –1.571, p = 0.043) and 02:00 (β = –1.763, p = 0.031) (Figure 2B). Sleep fragmentation was reduced by melatonin-suppressing light at 12:00 (β = –0.002, p = 0.039) and 13:00 (β = –0.003, p = 0.014) (Figure 2C).

**Figure 2.**
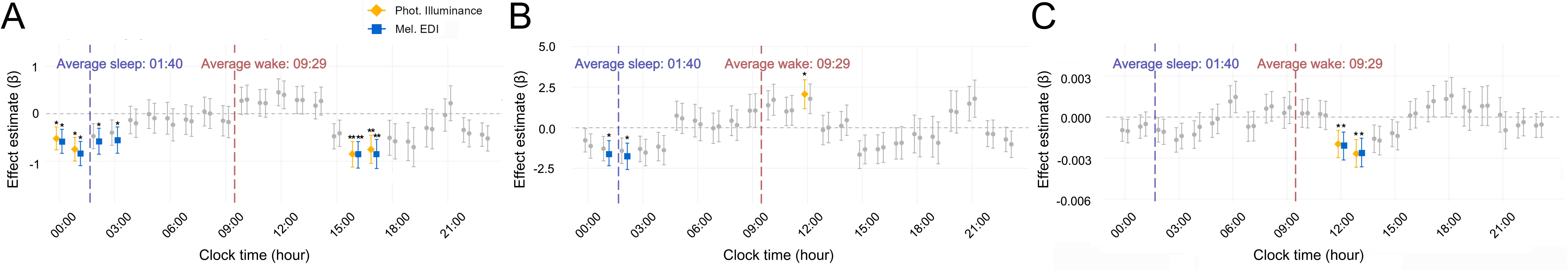
Hourly Effects of Melanopic and Photopic Light Exposure on Sleep Parameters. For all participants, (A) sleep quality Score, (B) deep sleep duration, and (C) sleep fragmentation. Yellow markers indicate significant effects of photopic illuminance; Blue markers indicate significant effects of Melanopic Equivalent Daylight Illuminance. Error bars represent ±1 standard error (SE) of the beta estimate. Vertical dashed lines mark average sleep onset and wake times to aid temporal interpretation.

### Chronotype-Specific Effects of Light Exposure on Sleep Parameters

In nET individuals, higher melanopic EDI during the nighttime period (00:00–06:00) significantly predicted lower sleep quality (β = –0.618, p < 0.001; Figure 3A). Hourly analysis further revealed that exposure at 01:00 (β = –0.804, p = 0.018) and 03:00 (β = –1.035, p = 0.015; Figure 4A) significantly predicted reductions in sleep quality, while elevated late-afternoon exposure at 15:00 (β = –0.767, p = 0.033) and 16:00 (β = –1.072, p = 0.011; Figure 4A) also exerted detrimental effects. In contrast, greater morning exposure at 10:00 significantly predicted improved sleep quality (β = 1.224, p = 0.002).

**Figure 3.**
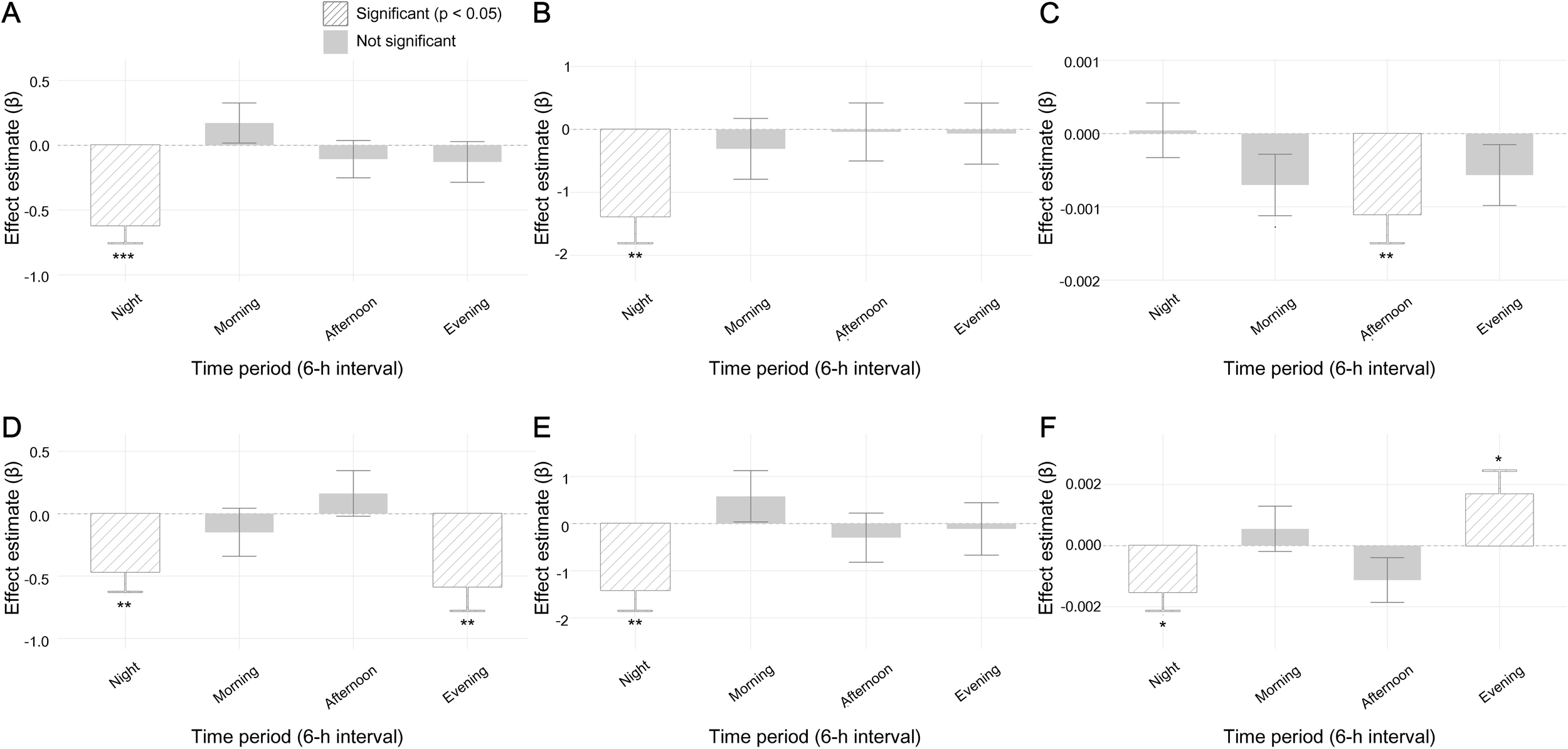
Chronotype-specific effects of light exposure on sleep parameters (6-hour interval). Results for non- Evening Types (A–C) and Evening Types (D–F) are shown separately. Estimates from linear mixed-effects models are presented for Night (00–06), Morning (06–12), Afternoon (12–18), and Evening (18–24). Panels display (A, D) sleep quality score, (B, E) deep sleep duration, and (C, F) sleep fragmentation. Bars represent effect estimates ±1 SE.

**Figure 4.**
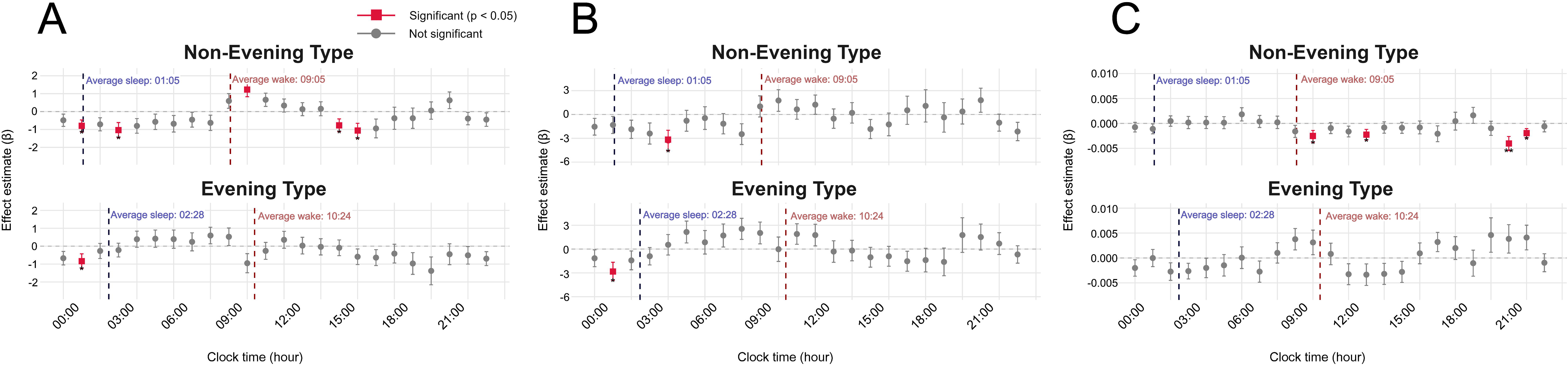
Hourly effects of light exposure on sleep parameters for Non-Evening Type (upper panels) and Evening Type (lower panels). (A) shows sleep quality score, (B) deep sleep duration, and (C) sleep fragmentation. Error bars represent ±1 standard error (SE) of the beta estimate.

For the same group, regression models indicated that greater nighttime light exposure significantly predicted shorter deep sleep duration (β = –1.384, p = 0.001; Figure 3B), with hourly analysis confirming a marked reduction at 04:00 (β = –3.277, p = 0.012; Figure 4B). Light exposure further predicted reduced sleep fragmentation overall (β = –0.001, p = 0.005; Figure 3C), with significant effects observed at 10:00 (β = –0.003, p = 0.031), 13:00 (β = –0.002, p = 0.036), 21:00 (β = –0.004, p = 0.005), and 22:00 (β = –0.002, p = 0.038; Figure 4C).

In ET individuals, regression models revealed that greater light exposure significantly predicted poorer sleep quality during both the night (00:00–06:00; β = –0.468, p = 0.003) and evening (18:00–24:00; β = –0.586, p = 0.002; Figure 3D). At the hourly level, only exposure at 01:00 significantly predicted lower quality (β = –0.837, p = 0.043; Figure 4A), while no significant associations were observed for morning or afternoon exposure. Deep sleep duration was also significantly reduced by greater nighttime light (β = –1.416, p = 0.001; Figure 3E), with hourly analysis confirming a decline at 01:00 (β = –2.798, p = 0.016; Figure 4E). For sleep fragmentation, the 6- hour model indicated significant associations with greater exposure at night (β = –0.002, p = 0.012) and in the evening (β = 0.002, p = 0.027), although these associations were not replicated in the hourly data, suggesting that broader temporal trends were more effectively captured at the aggregated level. Extended analyses across both temporal resolutions and chronotype groups are presented in Supplementary Figures S1–S6.

## Discussion

This study demonstrated that melanopic EDI was a more reliable and biologically meaningful predictor of sleep outcomes than conventional photopic illuminance under real-world conditions. Across all time windows, incorporating melanopic EDI into regression models consistently increased explanatory power for sleep quality, deep sleep duration, and sleep fragmentation. Unlike photopic illuminance, which reflects cone-mediated brightness perception, melanopic EDI captures the short-wavelength input to ipRGCs that regulate circadian and sleep processes [10, 55, 56]. This enhanced sensitivity to the blue-light spectrum likely accounts for its superior ability to predict various sleep parameters compared to conventional photopic measures, even under real-world lighting conditions. These findings provide field-based validation for melanopic EDI as a physiologically relevant light metric.

A clear distinction between melanopic and photopic measures also warrants emphasis. Within the CIE S 026 framework, melanopic EDI was proposed as a standardized metric for non-visual light responses [36]. While photopic illuminance adequately captures visual brightness, it frequently fails to predict physiological outcomes such as melatonin suppression, circadian phase shifts, or sleep disruption. The present results demonstrate that melanopic EDI, but not photopic illuminance, consistently predicted sleep outcomes across multiple domains. This provides strong support for adopting melanopic metrics in sleep and circadian research. The differential associations observed across sleep quality, deep sleep, and fragmentation may reflect the distinct physiological pathways these parameters index [57]. Melanopic-sensitive ipRGC signaling influences both circadian and direct sleep-regulatory processes, but the relative contribution of these mechanisms likely differs across sleep domains.

Nighttime exposure was associated with impaired sleep quality in both evening and non-evening types. This result extends well-established laboratory findings, where nocturnal or late-night light exposure suppresses melatonin secretion, delays circadian phase, and reduces sleep efficiency [58]. The present study demonstrates that these detrimental effects persist in naturalistic environments with variable exposure patterns, thus reinforcing the generalizability of prior experimental work. By showing that nighttime light degrades sleep across chronotypes under real-world conditions, this study underscores the importance of minimizing late-night exposure to melanopic-rich light.

Morning exposure exerted beneficial effects only in nET. Specifically, higher light levels at 10:00 were associated with improved sleep quality and reduced fragmentation. This is consistent with evidence showing that bright morning light advances circadian phase and stabilizes sleep in healthy and clinical populations [59]. In contrast, ET did not experience similar benefits, suggesting that morning light may not uniformly improve sleep across chronotypes. These results highlight the need for targeted light strategies, where nET individuals may derive more advantage from morning exposure interventions.

Evening light exposure impaired sleep specifically in ET participants. Higher light levels between 18:00 and 24:00 predicted lower sleep quality and greater fragmentation, effects not observed in nET. This vulnerability likely reflects the delayed circadian phase characteristic of ET, where evening light coincides with the pre-sleep period and further delays circadian timing. These findings align with prior evidence that late-day light disproportionately disrupts evening chronotypes and emphasize the necessity of stricter evening light control for ET individuals [60].

The implications of these findings extend to public health, clinical practice, and environmental design. Public recommendations should move beyond brightness alone to account for spectral composition and timing of light. Strategies may include reducing evening exposure to melanopic-rich light, while promoting morning exposure for nET individuals. In clinical practice, light therapy for insomnia, depression, or circadian rhythm sleep disorders may be optimized by integrating melanopic sensitivity and chronotype into treatment protocols. In applied contexts, smart environments and wearable technologies incorporating melanopic-sensitive monitoring could facilitate personalized circadian health strategies.

This study has several limitations. The sample consisted primarily of young adults, limiting generalizability to older or clinical populations. Sleep was assessed with a consumer wearable device rather than polysomnography, which may underestimate micro-arousals and subtle aspects of sleep architecture [58–60]. Light exposure was analyzed using fixed clock-time intervals rather than circadian phase markers such as dim-light melatonin onset (DLMO) [64], which may have led to some degree of phase misclassification.

In addition, although the light sensor provided reliable estimates of near-eye illumination, its collar placement may not fully capture corneal exposure under dynamic head movements or variable clothing conditions. Minor underestimation has been reported in high-luminance environments; however, this bias remains stable across intensity levels, supporting the sensor’s reliability for comparative analyses [38]. Similarly, the Fitbit Versa 4 may slightly underestimate total sleep duration and overestimate wake after sleep onset relative to polysomnography. Nevertheless, these deviations are systematic and consistent across participants, supporting its validity for detecting intra-individual variations [39, 40].

Despite these limitations, several strengths should be noted. Continuous near-eye monitoring of melanopic EDI enabled biologically relevant estimation of light exposure in real-world settings. The use of both hourly and 6- hour analyses allowed detection of temporal patterns that are often overlooked in broader time bins. The ecological validity of real-world data collection enhances the translational significance of the findings, and explicit inclusion of chronotype as a moderator provided novel insight into inter-individual variability in light sensitivity. Together, these features strengthen the robustness of the results and support their application to chronotype-specific light management strategies.

In conclusion, this study clarifies the relationship between light exposure and sleep by integrating melanopic metrics with chronotype-specific analyses in real-world settings. The results highlight that light evaluation should consider spectral composition, timing, and alignment with individual circadian rhythms. These insights provide a practical foundation for developing chronotype-tailored light hygiene strategies to improve sleep and circadian health.

## Author contributions

**Eunji Hwang:** Conceptualization, Methodology, Formal analysis, Investigation, Data curation, Visualization, Writing – original draft, Writing – review & editing. **Hyeonjin Kim:** Conceptualization, Supervision, Writing – review & editing. **Hahyun Lee:** Writing – review & editing. **Hyunwoo Nam:** Project administration, Funding acquisition, Resources, Supervision, Writing – review & editing. **Jun-Young Lee:** Conceptualization, Methodology, Supervision, Project administration, Funding acquisition, Resources, Investigation, Visualization, Writing – review & editing.

## Data Availability Statement

The data supporting the findings of this study are available from the corresponding author upon reasonable request.

