## Supplementary Figures for "Effects of Melanopic Equivalent Daylight Illuminance on Sleep Regulation and Chronotype-Specific Responses in Young Adults"

### Supplementary Material

**Eunji Hwang[1,2], Hyeonjin Kim[2], Hahyun Lee[2,3], Hyunwoo Nam[4,5,*], Jun-Young Lee[1,2,3,6,*]**

[1]Department of Medical Device Development, Seoul National University College of Medicine, Seoul, Republic of Korea

[2]Department of Psychiatry, Seoul Metropolitan Government Seoul National University Boramae Medical Center and Seoul National University College of Medicine, Seoul, Republic of Korea

[3]Interdisciplinary Program in Cognitive Science, Seoul National University, Seoul, Republic of Korea

[4]Department of Neurology, Seoul Metropolitan Government Seoul National University Boramae Medical Center and Seoul National University College of Medicine, Seoul, Republic of Korea

[5]Department of Neurology, Seoul National University College of Medicine, Seoul, Republic of Korea

[6]Department of Psychiatry, Seoul National University College of Medicine, Seoul, Republic of Korea

***Corresponding authors:**

Hyunwoo Nam,.

Jun-Young Lee,.

#### Supplemental Figures

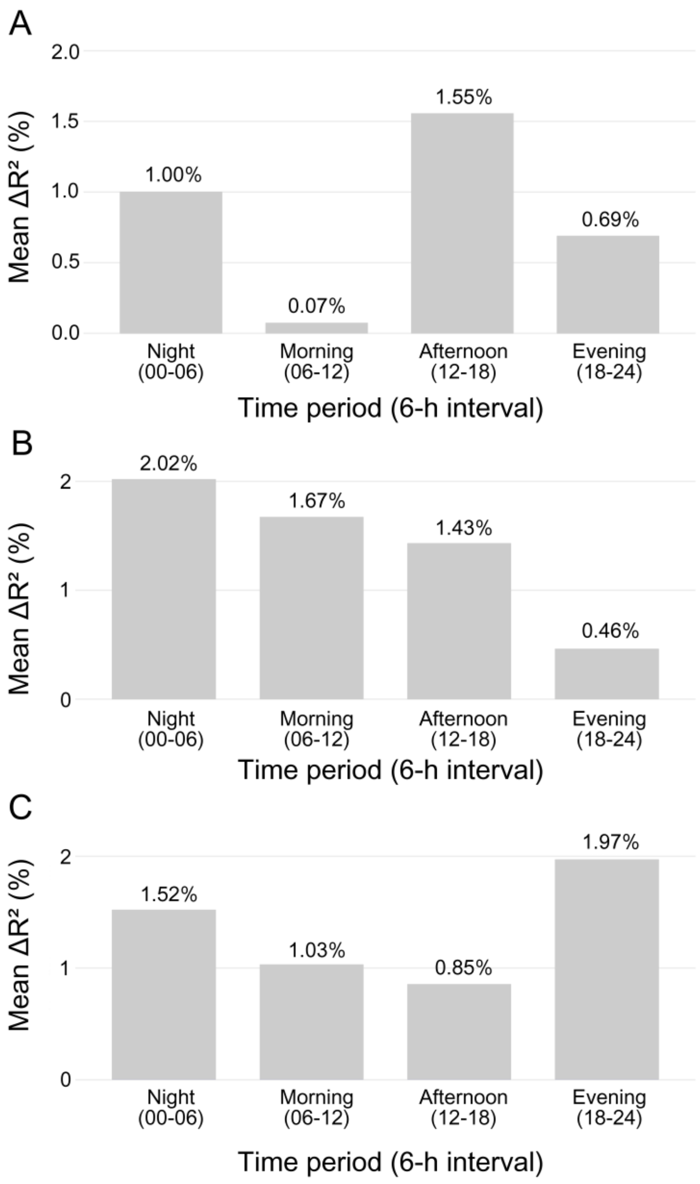


**Figure S1.** Mean $R^2$ improvement (6-hour bins) for Sleep Quality. Mean improvement in model $R^2$ (%) when melanopic equivalent daylight illuminance was added to photopic illuminance across four 6-hour periods (night 00–06 h, morning 06–12 h, afternoon 12–18 h, evening 18–24 h). Analyses were controlled for age, sex, employment status, and education. Panels A–C present results for all participants (n = 59), non-evening type (n = 38), and evening type (n = 21), respectively.

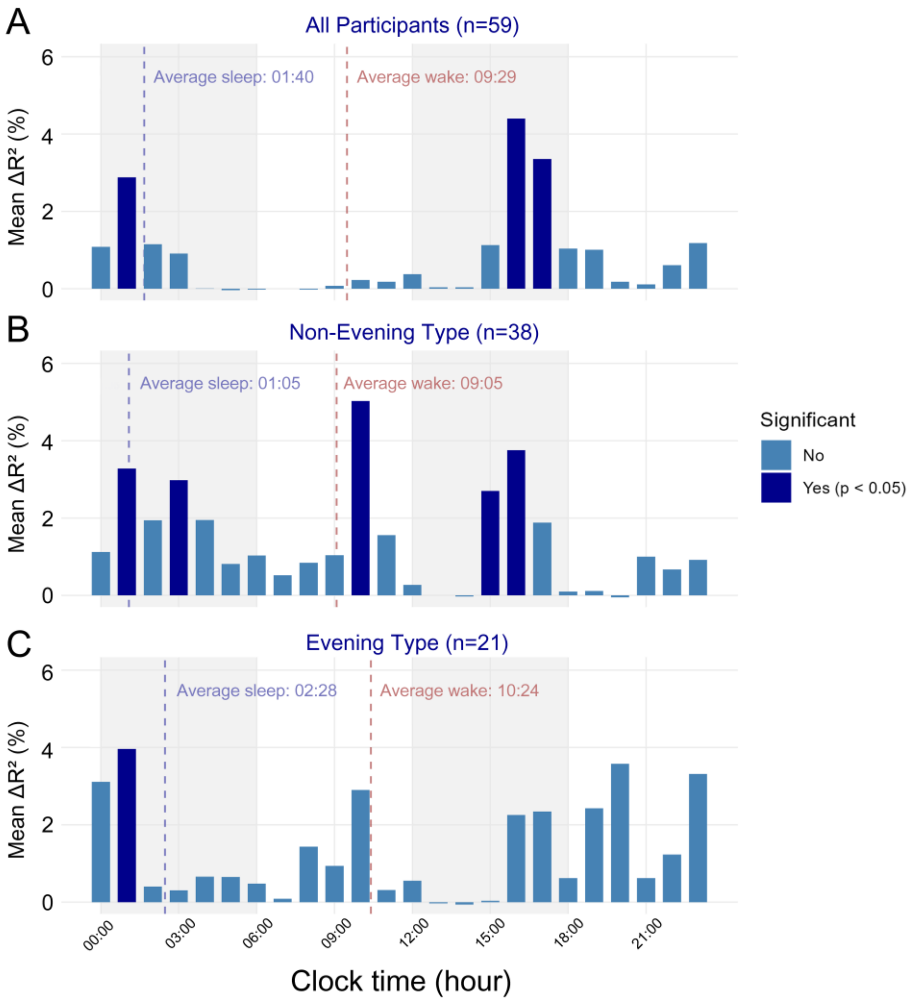


**Figure S2.** Hourly $R^2$ improvement for Sleep Quality. Hourly improvement in model $R^2$ (%) when melanopic equivalent daylight illuminance was included in addition to photopic illuminance. Panels A–C represent all participants, non-evening type, and evening type, respectively. Vertical dashed lines indicate the mean sleep and wake times for each group. The average sleep onset and wake times were 01:40 h and 09:29 h for all participants, 01:05 h and 09:05 h for the non-evening type, and 02:28 h and 10:24 h for the evening type.

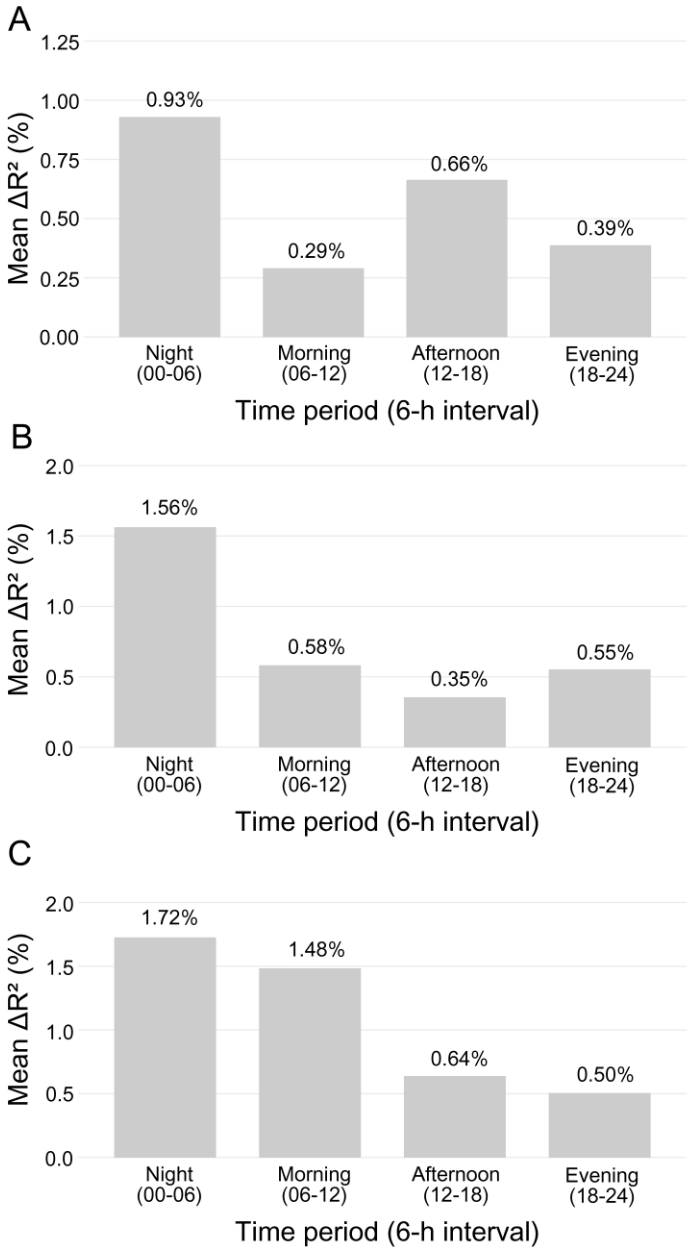


**Figure S3.** Mean $R^2$ improvement (6-hour bins) for Deep Sleep Duration. Mean improvement in model $R^2$ (%) for deep sleep duration when melanopic equivalent daylight illuminance was added to photopic illuminance. Analyses were adjusted for age, sex, employment status, and education. Panels A–C correspond to all participants, non-evening type, and evening type, respectively.

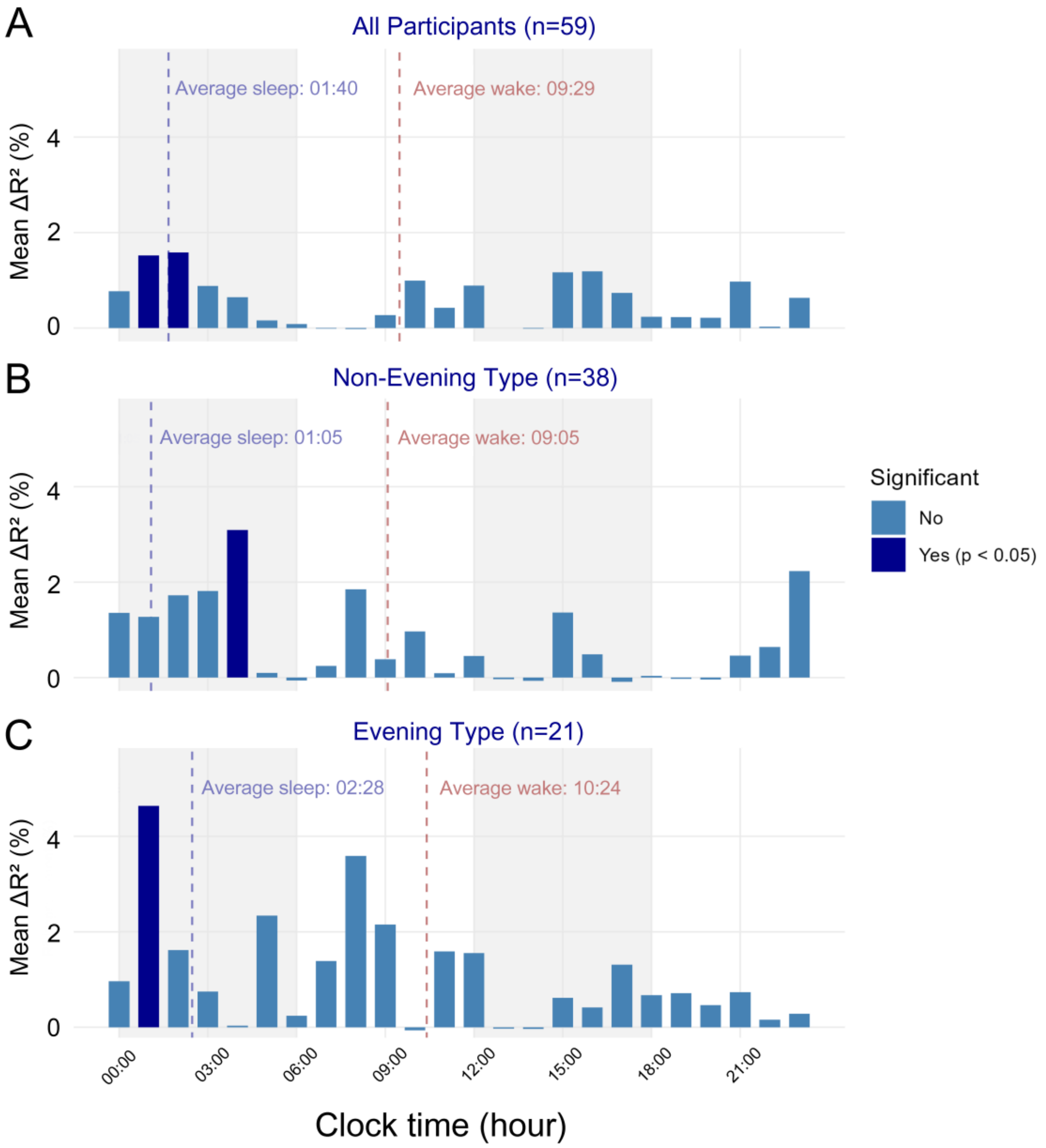


**Figure S4.** Hourly $R^2$ improvement for Deep Sleep Duration. Hourly variance improvement (%) in deep sleep duration predicted by melanopic equivalent daylight illuminance beyond photopic illuminance. Panels A–C correspond to all participants, non-evening type, and evening type. Vertical dashed lines indicate the mean sleep and wake times for each group. The average sleep onset and wake times were 01:40 h and 09:29 h for all participants, 01:05 h and 09:05 h for the non-evening type, and 02:28 h and 10:24 h for the evening type.

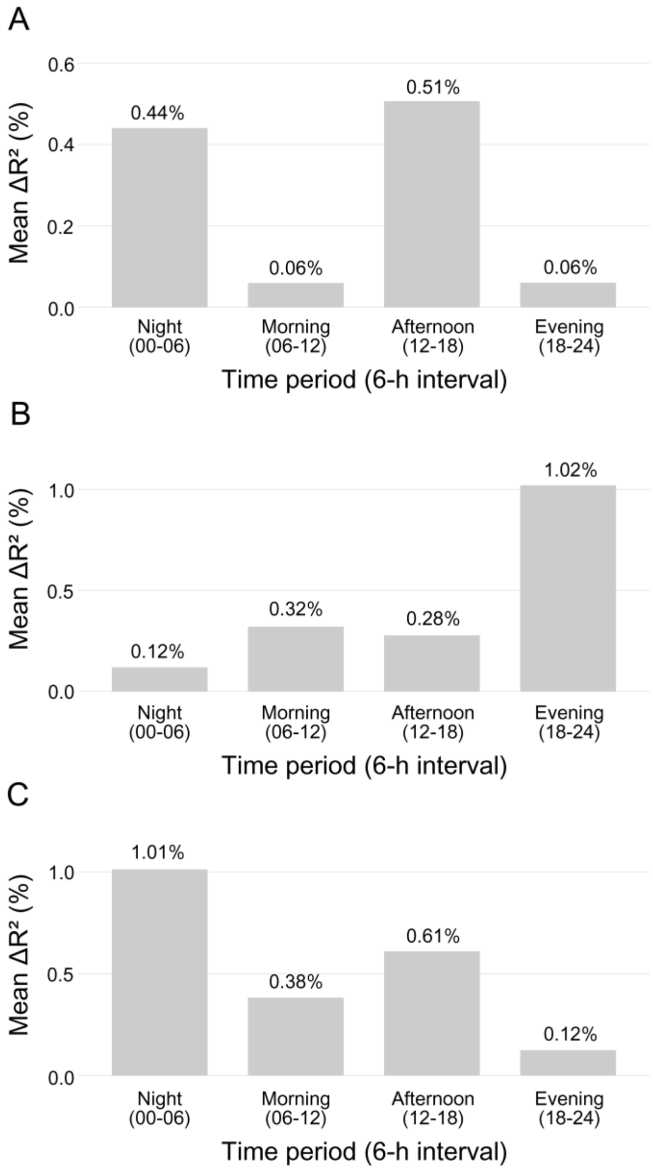


**Figure S5.** Mean $R^2$ improvement (6-hour bins) for Sleep Fragmentation. Mean improvement in model $R^2$ (%) when melanopic equivalent daylight illuminance was added to photopic illuminance models predicting sleep fragmentation. Analyses were controlled for age, sex, employment status, and education. Panels A–C present results for all participants, non-evening type, and evening type, respectively.

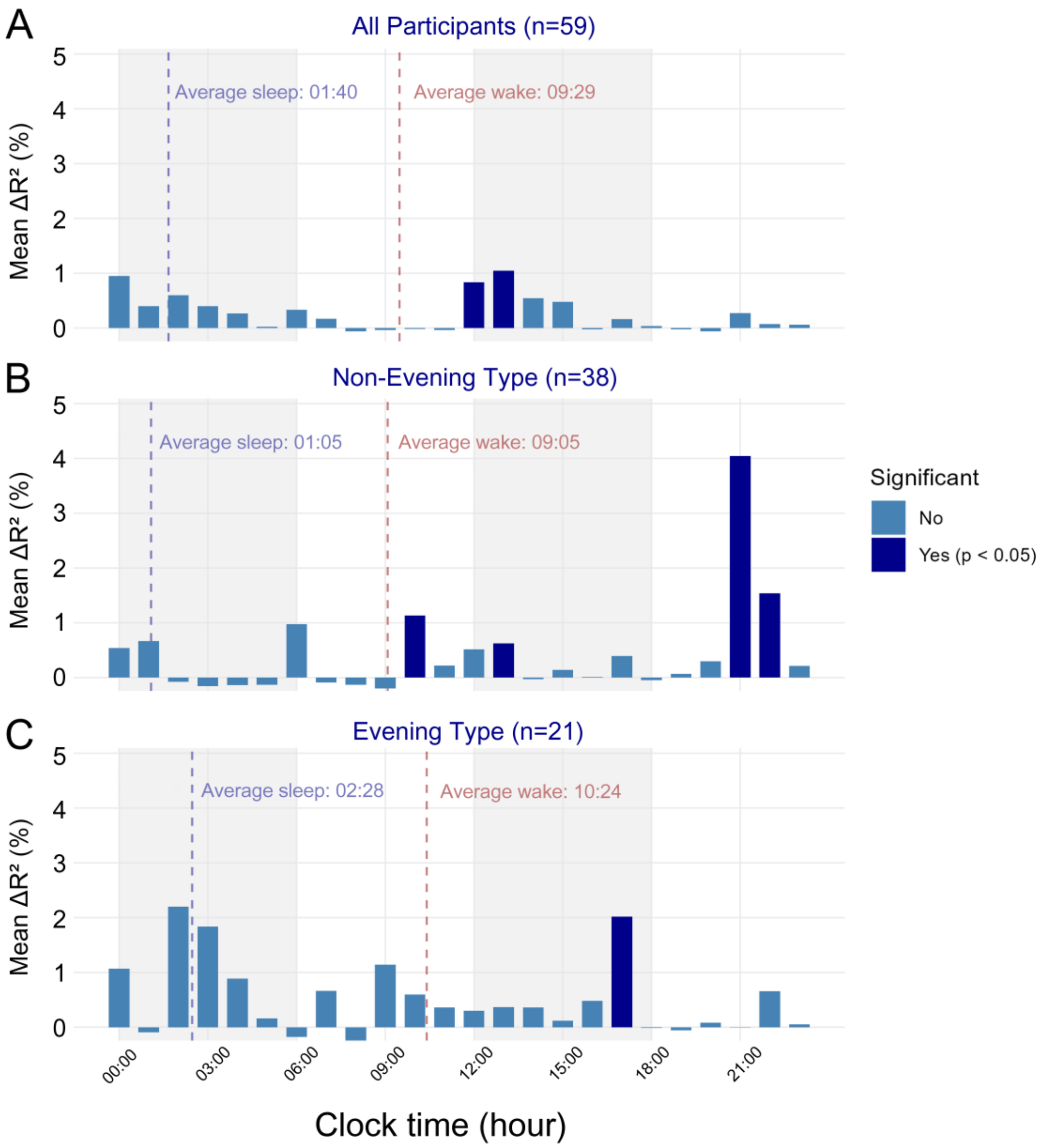


**Figure S6.** Hourly $R^2$ improvement for Sleep Fragmentation. Hourly improvement in model $R^2$ (%) for sleep fragmentation explained by melanopic equivalent daylight illuminance beyond photopic illuminance. Panels A–C show results for all participants, non-evening type, and evening type. The average sleep onset and wake times were 01:40 h and 09:29 h for all participants, 01:05 h and 09:05 h for the non-evening type, and 02:28 h and 10:24 h for the evening type. Vertical dashed lines indicate the mean sleep and wake times for each group.
